# The Effectiveness of Targeted Quarantine for Minimising Impact of COVID-19

**DOI:** 10.1101/2020.04.01.20049692

**Authors:** Alastair Jamieson-Lane, Eric Cytrnbaum

**Affiliations:** Institut für Chemie und Biologie des Meeres, Carl von Ossietzky Universität Oldenburg, Germany; Department of Mathematics, University of British Columbia, Canada

## Abstract

We model the extent to which age targeted quarantine can be used to reduce ICU admissions caused by novel coronavirus COVID-19. Using demographic data from New Zealand, we demonstrate that lowering the age threshold for quarantine to 50 years of age reduces ICU admissions drastically, and show that for sufficiently strict isolation protocols, isolating one third of the countries population for a total of 6 months is sufficient to avoid overwhelming ICU capacity throughout the entire course of the epidemic. Similar results are expected to hold for other countries, though some minor adaption will be required based on local age demographics and hospital facilities.

## 1 Introduction

COVID-19, initially observed/detected in Hubei province of China during December 2019, has since spread to all but a handful countries, causing (as of the time of writing) an estimated 855,000 infections and 42,000 deaths ([8], March 31st). COVID-19 has a basic reproductive number, *R*_0_, currently estimated in the region of 2.5 - 3 [5]. Social distance and general quarantine measures can reduce *R*_0_ temporarily, but not permanently. For *R*_0_ = 3, left unchecked COVID-19 can be expected to infect more than 90% of the population, with 30% of the population infected at the epidemic peak. Even with significant quarantine measures in place the population will not reach “herd immunity” to this virus until 2/3 of the population has gained resistance-either through vaccination, or infection and subsequent recovery.

In order to place these numbers in a concrete context, a recent survey in New Zealand indicated that the country had a total of 520 ventilator machines [7]. Given the country’s demographics (see table 1), and current estimates of ICU risk per age cohort, the country would expect to see at minimum 66,000 ICU patients over the course of an epidemic. Thus, assuming that severe cases require 14 days in ICU, treatment of all patients would require over 900,000 ventilator days – amounting to five years of continuous use of all ventilators. This is 15 times more demand than could be accommodated in the expected 4 month span of an unmitigated epidemic. The details of this calculation may vary from country to county, but the final conclusion is ubiquitous: hospitals are not prepared for this disease.

**Table 1:** Here we provide demographic data for New Zealand [2], along with risk of ICU admission per infection for each age group [1]. Finally we give the *expected* number of ICU admissions, assuming 2*/*3^*rd*^s of each age category becomes infected over the course of the epidemic-the minimum required to reach herd immunity.

| Age | Population | ICU risk | Expected ICU admissions |
| --- | --- | --- | --- |
| 0-9 | 615,364 | 0.005% | 20 |
| 10-19 | 606,025 | 0.015% | 61 |
| 20-29 | 664,742 | 0.06% | 266 |
| 30-39 | 611,029 | 0.16% | 651 |
| 40-49 | 613,469 | 0.31% | 1263 |
| 50-59 | 611,028 | 1.2% | 5,069 |
| 60-69 | 488,835 | 4.5% | 14,823 |
| 70-79 | 317,385 | 10.5% | 22,212 |
| 80+ | 169,555 | 19.4% | 21,879 |

Efforts to “flatten the curve” will need to reduce the epidemic peak not merely by a factor of two, but instead by an order of magnitude or more. Even in the most optimistic scenarios, for the most well equipped countries, such efforts must be maintained for years on end.

Societal lockdown may be effective at eradicating COVID-19 locally, but when lockdown is complete a large susceptible population will remain; if the virus is later re-introduced, as expected in our globalized and interconnected world, a new epidemic is likely to occur. While buying time allows for manufacturing of new medical equipment, and further scientific investigations, such efforts can not be maintained indefinitely, and even optimistic predictions predict a vaccine will take no less than one full year to develop. For this reason it proves necessary to discuss not just what quarantine measures are needed, but also how society might return to normal, and over what time frame this can be achieved.

## 2 Targeted Quarantine and Release

As has been observed in South Korea[4], death rate is tightly correlated with age. While deaths in younger age categories are observed, a recent report from Italy[3] indicates that the vast majority of deaths occur in patients with known pathologies. It should thus be possible to predict who is most at risk with high fidelity. By controlling *which* 2/3rds of the population become infected we may avoid overtaxing the healthcare system and thus minimise mortality.

We consider a strategy in which initially all of society (bar essential service workers) are locked down for two to four weeks so as to brake the initial uncontrolled epidemic spread. At the end of this time, rather then reduce quarantine measures uniformly over all society, a government could instead engage in a program of ‘targeted quarantine’; those individuals with no risk factors would be encouraged to return to work and socialize, while at risk individuals would be asked to remain at home under strict quarantine. The infection would be allowed to pass through the large, low risk population, resulting in a small number of ICU admissions due to occasional unexpected complications. Once the epidemic has passed through the low risk population, quarantine measures can be lowered in stages, with each stage exposing successively higher risk populations. Despite the increased risk on the individual level, the reduced susceptible population for each subsequent stage would lead to a natural ‘flattening of the curve’, and a smaller fraction of these later populations becoming infected. Eventually herd immunity would be achieved, and isolation measures can be safely dropped for the entire population. Figure 1 gives a schematic representation of this approach.

**Figure 1:**
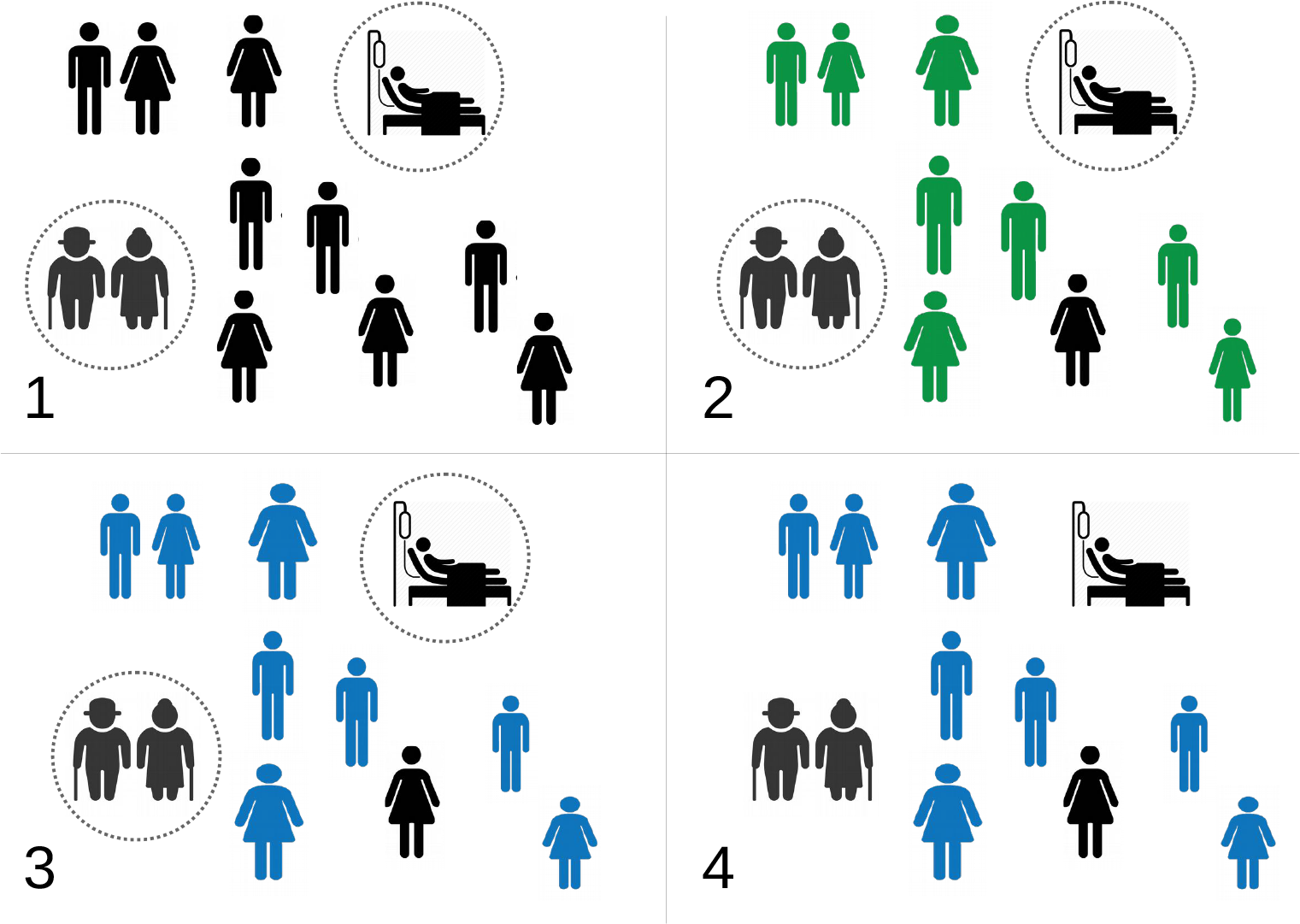
A schematic illustration of targeted quarantine. Susceptible population in black, infectious population in green, and recovered population in blue. 1) we isolate at risk populations as quickly, and as strictly as possible. 2) COVID-19 is allow to spread through the large, low risk population. This leads to a small number of hospitalizations, which the hospital system is able to cope with. 3) The low risk population recovers, and herd immunity is achieved. 4) Quarantine is (cautiously) removed, and high risk individuals are able to return to normal social activity.

Similar strategies were alluded to by Ferguson *et al* [1], who advised social distancing for those over the age of 70, and investigated explicitly by Chikina & Pegden [6], who completed a comprehensive sensitivity analysis suggesting a significant reduction in mortality for a wide range of parameters. Both papers however, proposed a fixed age of division (70 and 65 years of age, respectively), and explored what might be considered reasonable parameter ranges in terms of levels of isolation possible, predicting, even in their most optimistic estimates, hundreds of thousands of fatalities. There is, however, no reason to restrict the targeted quarantine threshold to 65 or 70 years of age, and, as we shall show, lowering this age threshold to 50 can reduce fatalities substantially.

## 3 The Model

In order to illustrate the general approach described above, we consider a simple SIR type model, in which our population is divided both by disease status (Susceptible, Infected, Recovered), and by age cohort. Here we lump people by decade, as this is the granularity that fatality rates and ICU admission data are reported. We assume fixed recovery rate *γ* = 0.1 days^*−* 1^, and a fixed infection rate *β* = 0.25 days^*−* 1^. Interactions between age cohorts are governed by the contact matrix ***κ***(*t*). Entry *κ*_*i,j*_ gives the contact rate between age cohort *i* and age cohort *j*. This is the system parameter that can be changed through public policy and human behavior.

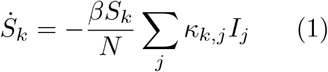

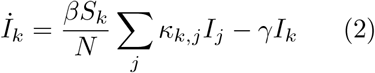

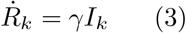

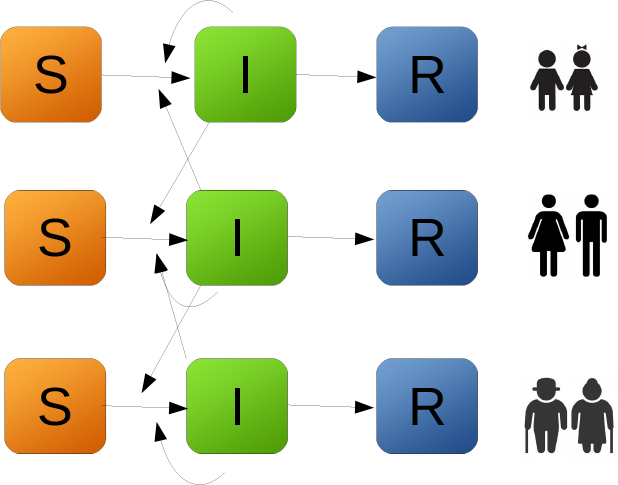

In order to simulate targeted quarantine measures, we select some threshold age *A*, and some time window (*t*_*s*_, *t*_*e*_) during which quarantine measures apply. We define two contact matrices, one for the quarantine period, and one for the non-quarantine period. These matrices take the form:

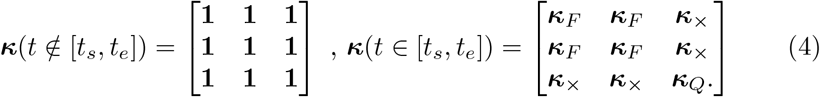

During quarantine, we assume a contact rate ***κ***_*i,j*_(*t*) = *κ*_*Q*_ = 0.1, for individuals within the quarantine (*i, j > A*), and ***κ***_*i,j*_(*t*) = *κ*_*×*_ = 0.1 across the quarantine barrier. Individuals outside of the quarantine group (*i, j ≤ A*) have contact rate ***κ***_*i,j*_(*t*) = *κ*_*F*_ = *N/N*_*nq*_ where *N*_*nq*_ is the total population *not* in quarantine (*N*_*q*_ gives the population in quarantine. Similarly *S*_*q*_ gives the susceptible population under quarantine, and *I*_*nq*_ gives the infectious population not under quarantine). This choice of *κ*_*F*_ approximately preserves the total number of interactions experienced by the out of quarantine population, by compensate for the loss of interaction caused by the removal of the quarantined population.

ICU demand at any given time is given by ∑*α*_*k*_*I*_*k*_, where here *α*_*k*_ is the probability of ICU admission for age category *k* (see table 1). Cumulative ICU bed demand is *α*_*k*_(*I*_*k*_ +*R*_*k*_). Full R code for all models is publicly available on Github ^1^. Readers are encouraged to adapt demographic and clinical parameters based on their own circumstances, and in response to further research.

## 4 Results

Simulation of the above system for a range of quarantine thresholds A results in ICU demand as depicted in Figure 2. Figure 3 summarizes peak ICU demand as a function of A. Peak ICU demand is minimized for a quarantined threshold of *A≈* 40, and total ICU demand is minimized for *A≈*50.

**Figure 2:**
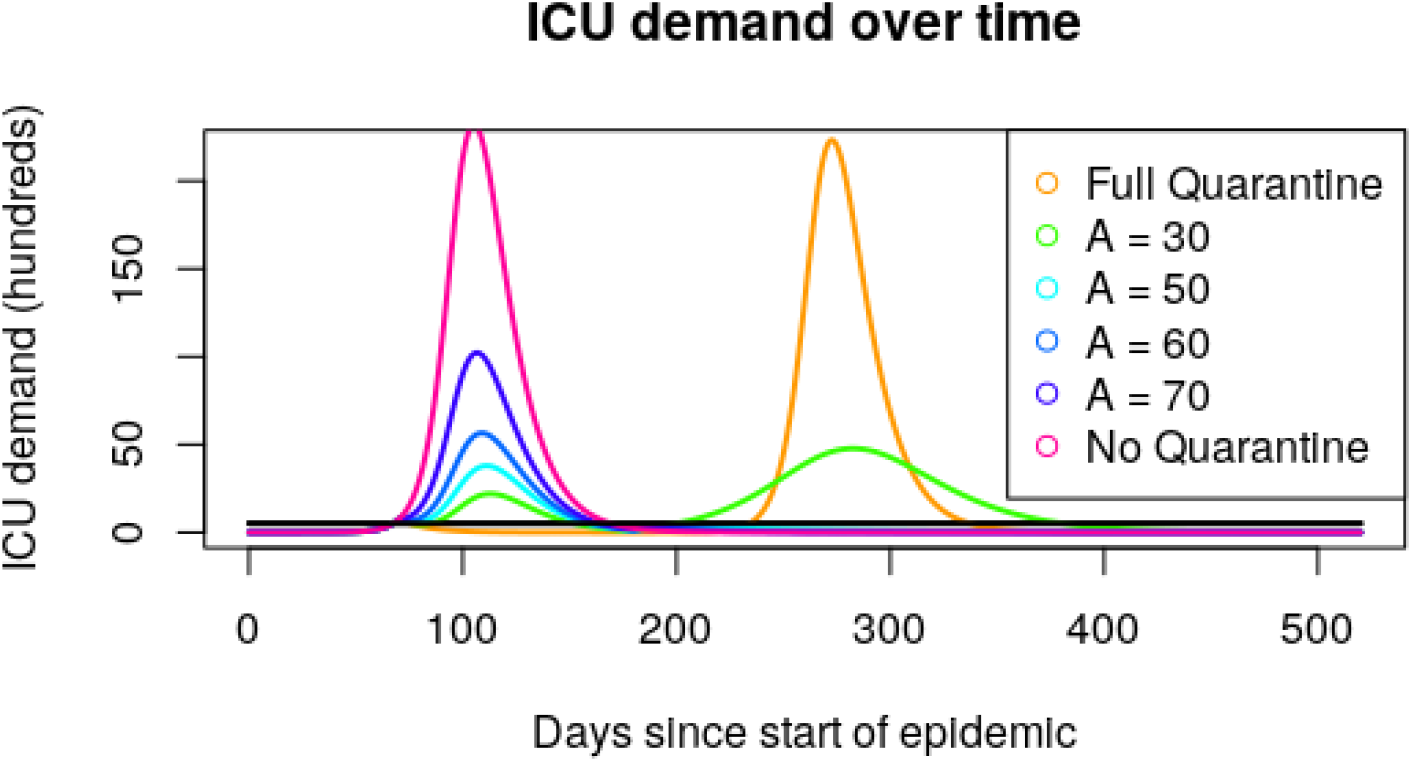
Epidemic curves assuming 110 days of quarantine (*t*_*s*_ = 70, *t*_*e*_ = 180), for a variety of age thresholds *A*. Total Ventilator numbers are given by the black line. Note that the results of quarantining everyone and quarantining no one are largely the same, with quarantine simply delaying the peak. An age cut off of 50 years (teal curve) captures a large enough portion of the population to achieve herd immunity, while reducing fatalities compared to cut offs at 60 or 70 years.

**Figure 3:**
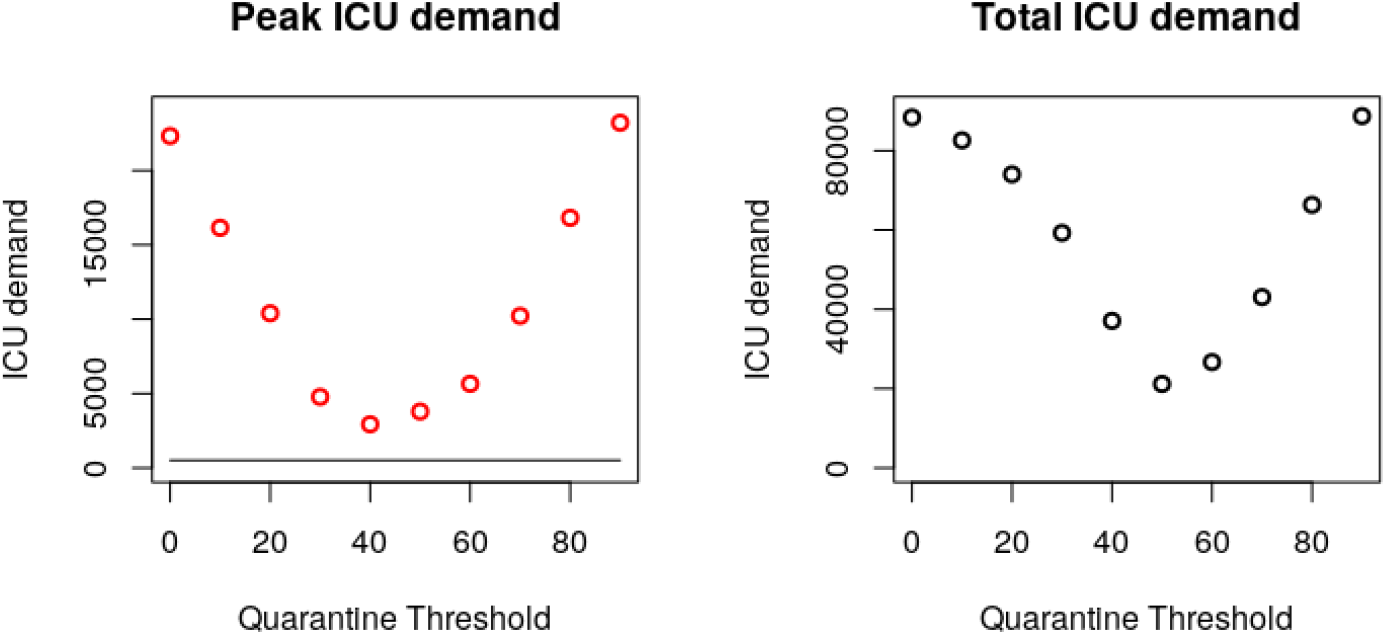
Depending on the placement of our quarantine threshold, both peak and total ICU admissions vary. Given that hospital facilities are rapidly over-whelmed in even the best case scenario above, total ICU demand is expected to be a close proxy for total fatalities. A threshold at 50 years of age minimizes this number.

In order to understand the importance of our various quarantine compliance parameters, it proves useful to determine the total number of infections due to “quarantine leakage”. This value is well approximated by

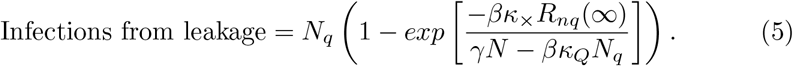

Derivation of this expression is given in the Appendix. Figure 4 verifies this expression for a variety of *κ*_*×*_ values by comparing equation 5 to the results of full scale SIR simulation.

**Figure 4:**
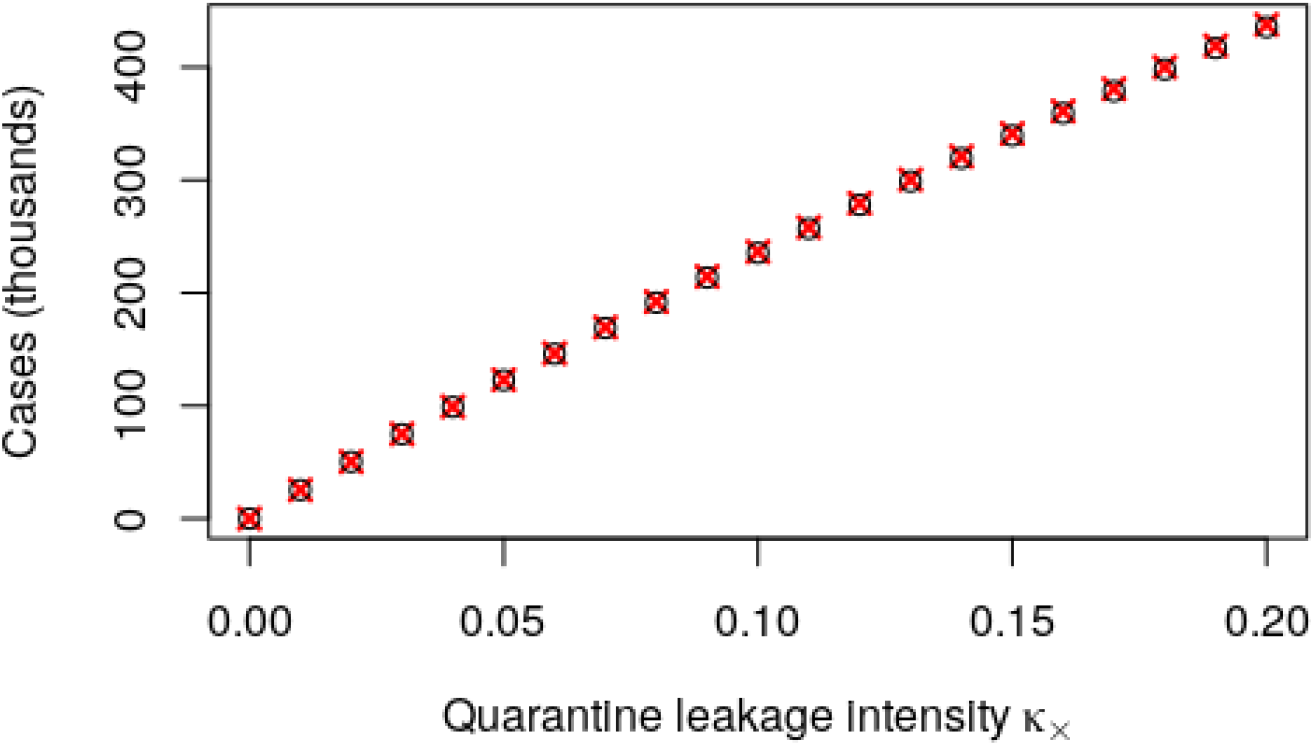
Total infections caused by quarantine leakage assuming *κ*_*Q*_ = 0.1. Black circles give the predictions based on full SIR dynamics, red crosses give the predictions of equation 5. Note that these calculations *only* include infections caused by imperfect quarantine, and ignore infections occurring either outside the quarantine population, or outside the quarantine time period.

While complicated at first glance, equation 5 provides two key insights: firstly, we observe that *all* reductions in cross quarantine contact rate, *κ*_*×*_, lead to a corresponding reduction in infections - halving *κ*_*×*_ will (approximately) halve the number of quarantined individuals getting infected. Secondly, we observe the importance of keeping *κ*_*Q*_ well below the epidemic threshold. So long as *γ » βκ*_*Q*_*N*_*q*_*/N*, quarantine measures are strict enough to prevent self-sustaining chains of infection in the quarantine population and *κ*_*Q*_ has only moderate impact. However, if the within quarantine contact rate *κ*_*Q*_ approaches the threshold *γ* = *βκ*_*Q*_*N*_*q*_*/N*, the number of infections increases *very* quickly. Hence while it is preferable to keep *κ*_*×*_ as small as possible, it is *critical* to keep *κ*_*Q*_ small *enough*. Once this has been achieved, efforts can be focused elsewhere.

So far we have seen how careful selection of age threshold can reduce ICU admission (figure 3), and also how reducing cross quarantine contact reduces infection (figure 4). This leads naturally to the question of “What is our best case scenario? How do we get there?”. In figure 3 we showed how a single stage quarantine approach can decrease mortality. In figure 5 we show that a two stage quarantine approach can do even better, under ideal circumstances bringing peak ICU demand below capacity. Given that ICU overflow translates almost directly to patient mortality, this is a *very* desirable goal.

**Figure 5:**
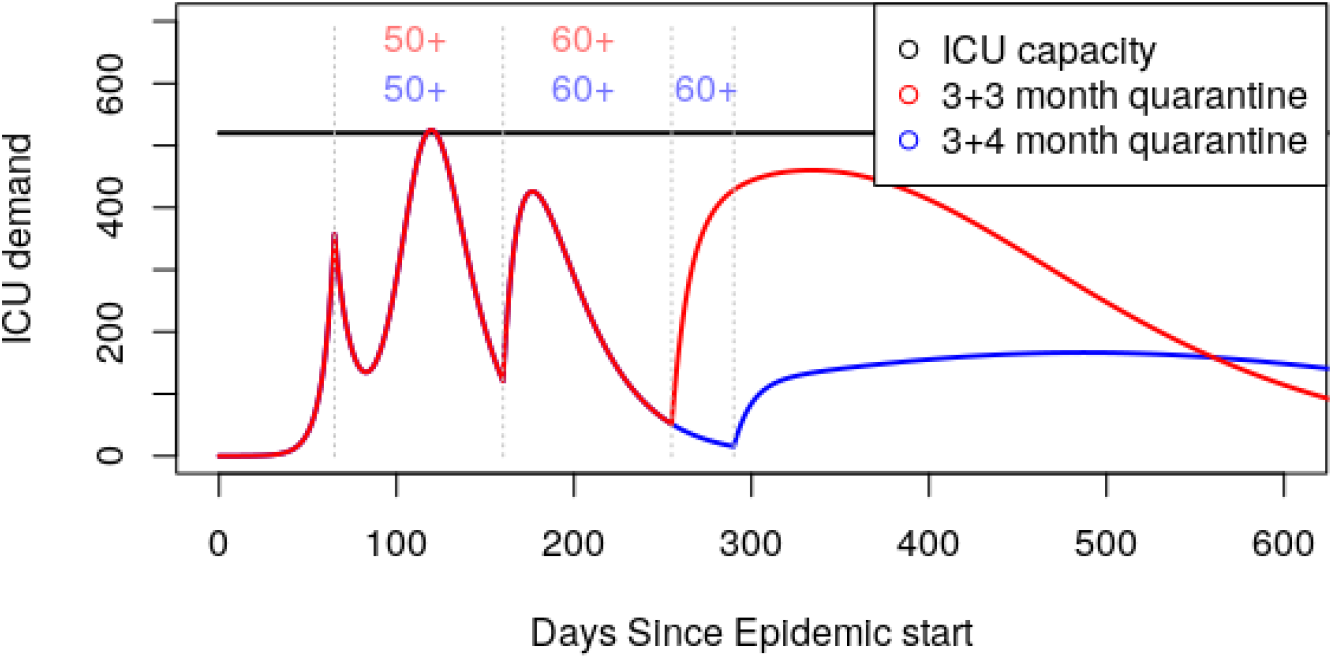
Assuming perfect quarantine between age groups (*κ*_*×*_ = 0), we are able to keep ICU demand below ICU capacity using a multi-stage release strategy. For the red curve, we assume quarantine of everyone over 50 years of age from day 65 to 160, and quarantine of everyone over 60 years of age from day 160 to 255 (that is to say, 3 month intervals for each stage of isolation). At no point is the under 50 population required to remain in quarantine, although moderate social distancing is required in the first 3 month window. Isolating only the 60+ cohort in the first 3 month window (as opposed to 50+) leads to overflowing ICU capacity by a factor of 5. An alternative strategy (blue), extends quarantine until day 290 (an extra month). This extended quarantine of the over 60 cohort reduces total ICU admissions from 17500 to 11400, and spreads these resulting admissions over a far wider time span.

## 5 Discussion

### 5.1 Logistics

Above we have implemented a variety of possible containment strategies through simply changing a few parameter values. Implementing such policies in real life will be significantly harder. A few of the most pressing logistical issues which need analysis are:

- Establishing food delivery infrastructure so that quarantined individuals can stay at home.
- Partitioning health care facilities so as to prevent mixing between quarantine groups.
- Communication and public education so that people understand both what is required for the approach to work, and how it benefits them.
- Rearranging our workforce, so as to account for the large number of senior staff who will no longer be available in person.
- Identifying new jobs that will need to be carried out, so as to maintain quarantine as effectively as possible.
- Investigation and community engagement with ‘de-mixing’ households, so that people in different quarantine groups are no longer living with one another. Such measures may be disruptive, but are likely necessary if we wish to minimize *κ*_*×*_, and would (for example) involve children staying with aunties and uncles, or family friends, rather than grandparents. Individuals in the quarantine group currently living alone might consider moving in with other quarantined persons, and university students would be required *not* to return to their parents households.

### 5.2 Assumptions to be Investigated

We have explored means of minimizing the death toll and burden on the medical system. This work is based on a number of assumptions. Each of these assumptions must be verified before any of the above plans are implemented:

- We have taken the ICU admission rates provided by Ferguson et al. as our baseline assumption[1]. If it is later found that the ICU admission risk amongst younger generations are higher than expected, it is likely that ICU capacity will be exceeded. Nonetheless, separation of age cohorts still leads to a net improvement in outcome, as it allows the medical system to deal with separate epidemic peaks as opposed to a single larger peak [6].
- We assumed that infection provides immunity, or at least resistance to COVID-19. If it is found that infection does not result in subsequent resistance, or resistance is short lived, then any protocol that depends on herd immunity will fail.
- We have made simplifying assumptions about community structure. It is likely that community structure (for example, retirement homes, where high risk individuals interact frequently with other high risk individuals) may increase the threshold required for herd immunity to be acquired.

### 5.3 Opportunities

There are a number of research avenues that may improve upon the outcomes we have described.

We have considered a simple age structured population; this was driven primarily by the availability of age structured ICU risk data. With better understanding of the underlying health conditions associated with severe COVID-19 outcomes, and better demographic data on the frequency of these risks, it will be possible to pre-isolate high risk individuals in our under 50 age cohort, and simultaneously, reduce quarantine requirements for individuals over 50 who have a clean bill of health.

We assumed a “predetermined” epidemic response strategy. Future developments in serological testing would allow more dynamic strategies to be adopted, based on weekly testing of antibody levels in the population; decisions on quarantine measures could then be made based on observed immunity levels in the population, rather than simply occurring at three month intervals. Such screening and feedback would greatly mitigate the various uncertainties in the model. Finally, although here we have considered age-targeted quarantine approaches as they apply to the population of New Zealand, this was merely for the sake of concreteness; nothing in the approach described is specific to that country. Readers are encouraged to make use of our code, and explore the dynamics given their own local demographic data.

## 6 Conclusions

While previous authors [1] [6] have alluded to and explored the use of age targeting, they have done so in the limited sense of quarantining retirees, and assumed moderate social distancing measures are implemented. Here we argue that this undersells the potential effectiveness of the targeted quarantine approach; any government willing to take the extraordinary measure of shutting down all of society should, at the very least, *consider* the significantly less drastic measure of quarantining one third of their population.

To the low risk population, our strategy offers a fast return to functional society, and, for those few who do get sick, access to medical facilities and staff who are dealing with a manageable number of cases. To those under isolation, targeted quarantine offers the possibility of avoiding COVID-19 altogether. By releasing a large portion of the population from lockdown after a small number of weeks, age-targeted quarantine balances the need to maintain a functional society and economy, with the limitations of hospital resources, and has the potential to significantly reduce casualties.

## Data Availability

Code used for simulations is available on github.

https://github.com/alastair-JL/COVID_strategy

## 7 Acknowledgements

Funding was provided by the School of Medicine and Health Science, Carl von Ossietzky Universität Oldenburg. This work is based on modelling done in collaboration with the Interventional Modelling Team at the Department of Medical Microbiology and Infection Prevention at the University Medical Center Groningen. Further advice and feedback was provided by the the Mathematical Biology team of the University of British Columbia. Special thanks to Corinna Glasner for guidance and editorial advice on an early version of this manuscript, and to Alejandra Herrera for some added clarity and final polish.

## 8 Appendix Analytic formula for the effects of quarantine leakage

In order to understand the effects of our various parameters, we wish to estimate the effects of ‘quarantine leakage’. While simulations determine such results quickly enough, determination of an analytic expression is useful for quickly judging the importance of each parameter. In order to determine such a formula, we proceed in three steps.

First, we calculate the number of infections a quarantined individual expected to receive from the non-quarantined population. This is equal to 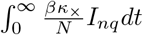 Because 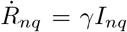 we find 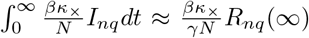. The value *R*_*nq*_(*∞*) is a function of *R*_0_, found by solving 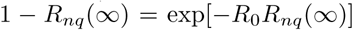. In the case *R*_0_ = 2.5, *R*_*nq*_(∞) = 0.89.

For our next step, we must account for the fact that infections that enter our quarantined population may then spread (especially important when *κ*_*×*_ *< κ*_*Q*_). Whenever 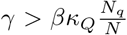, transmission is unable to sustain itself and this spread can be modeled as a subcritical branching process. Such a process has leads to an expected population of *γ/*(*γ − βκ*_*Q*_*N*_*q*_*/N*) infections before extinction-that is to say, each imported infection is expected to lead to *γ/*(*γ − βκ*_*Q*_*N*_*q*_*/N*) infections total.

Multiplying the results of the previous two steps gives 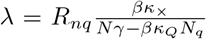, the total number of infections due to quarantine leakage that each quarantined individual *expects* to encounter. The actual number of infections a given individual receives is Poisson distributed, and the probability of receiving zero infections is thus given by *e*^*−λ*^.In total combining the above, we end up with

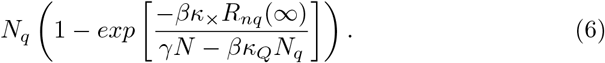

individual infected due to quarantine leakage. This is give as equation 5 in the main text.

## Footnotes

1 https://github.com/alastair-JL/COVID_strategy

